# An Integrated Platelet Index Derived From PCA Is Associated With Long-Term Atherothrombotic Risk in Acute Coronary Syndrome

**DOI:** 10.64898/2025.12.15.25342328

**Authors:** Yuefeng Wang, Jiayi Sun, Zekun Zhang, Ruiyue Yang, Jun Dong, Yuxuan Li, Xingyun Jiao, Yixiang Liu, Xingyu Chen, Wei Gong, Xue Yu

## Abstract

**Background:** Platelet morphological indices, such as mean platelet volume and distribution width, reflect platelet turnover and thrombo-inflammatory activity relevant to acute coronary syndrome (ACS), but their individual prognostic value remains inconsistent. This study aimed to derive an Integrated Platelet Index (IPI) using principal component analysis (PCA) to capture a multidimensional platelet morphological phenotype influenced by cardiometabolic and inflammatory stress and to examine its association with long-term major adverse cardiovascular and cerebrovascular events (MACCE) in ACS.

**Methods:** This prospective observational study included 1,467 ACS patients from the Beijing Hospital Atherosclerosis Study. Five routinely measured platelet indices—platelet count, mean platelet volume, platelet distribution width, plateletcrit, and platelet large-cell ratio—were integrated via PCA to construct the IPI. The primary endpoint was MACCE. Cox models, restricted cubic splines, Kaplan–Meier curves, and prespecified subgroup analyses assessed the prognostic relevance of the IPI and its consistency across metabolic and clinical strata.

**Results:** Over a median follow-up of 55.0 months, 141 patients (9.6%) experienced MACCE. Lower IPI values were associated with higher risk. Each unit increase in IPI was independently associated with a 27% lower MACCE risk after full adjustment (HR 0.73; 95% CI: 0.59–0.90). A significant L-shaped nonlinear relationship was observed, with steep risk reduction at lower IPI values. Event-free survival increased progressively across IPI tertiles. Associations remained consistent across cardiometabolic subgroups, with no significant interactions.

**Conclusions:** The PCA-derived IPI represents an integrated platelet morphological phenotype associated with long-term atherothrombotic risk in ACS. Its nonlinear behavior and robust performance across metabolic backgrounds support its potential as an accessible, phenotype-based marker for refining secondary prevention risk stratification.

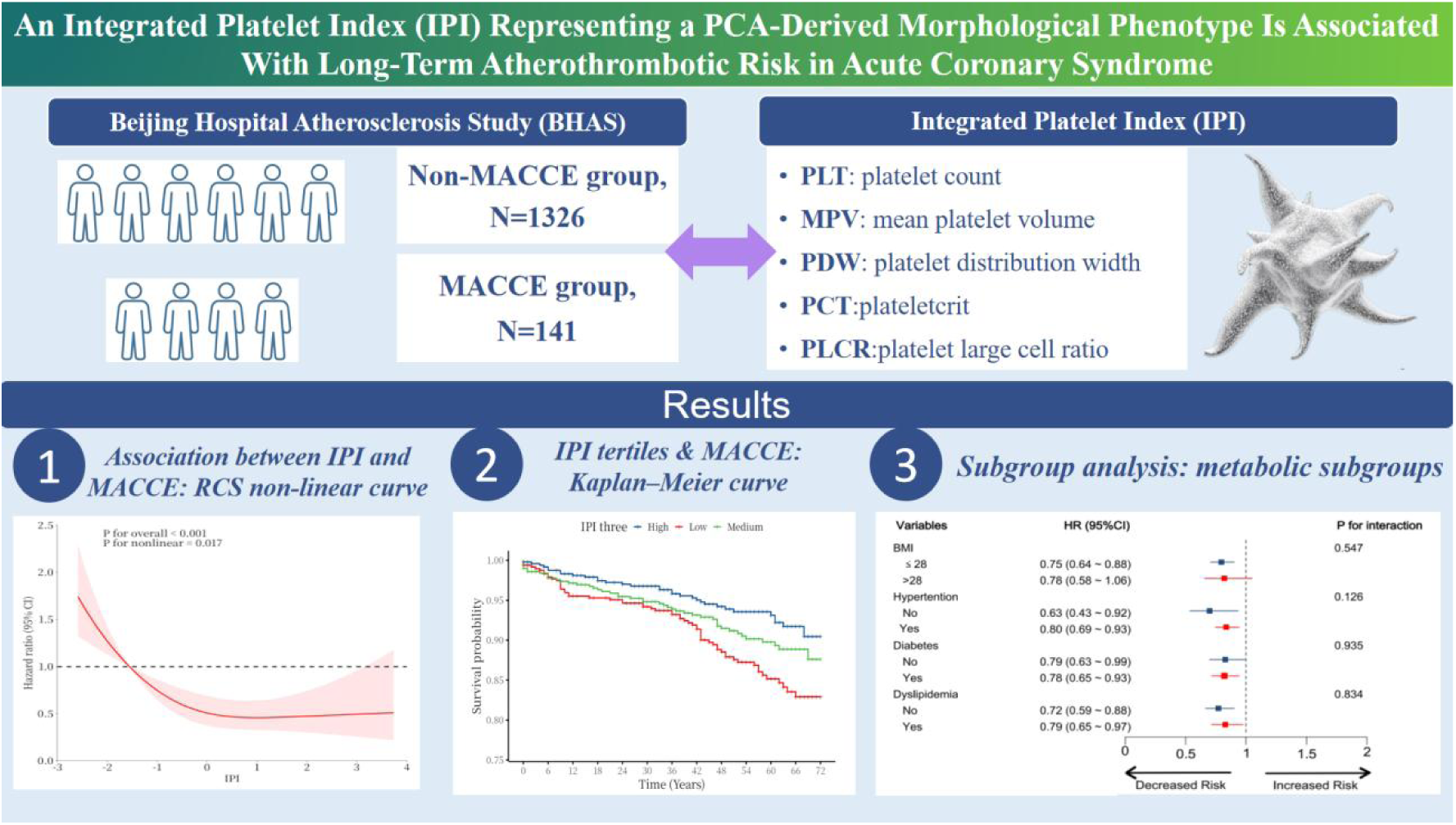

**What Are the Clinical Implications?:** Platelet morphology and turnover are influenced by cardiometabolic and inflammatory stress, yet routine platelet indices are rarely incorporated into ACS risk assessment because each parameter reflects only a single dimension of platelet biology. By integrating five commonly available platelet indices into a single PCA-derived morphological phenotype, the Integrated Platelet Index (IPI) provides a multidimensional representation of platelet alterations relevant to atherothrombotic risk.

In this study, lower IPI values consistently identified patients at higher long-term risk of MACCE, and the L-shaped nonlinear association suggests that relatively small reductions in this phenotype may reflect disproportionately adverse platelet remodeling. Because the IPI relies solely on automated complete blood count parameters, it is low-cost, universally available, and feasible for implementation in secondary prevention pathways, including in resource-limited settings.

Clinically, the IPI may improve risk stratification in ACS by capturing thrombo-inflammatory and metabolic influences not detected by traditional predictors. Its stability across cardiometabolic subgroups supports broad applicability. Future studies should determine whether serial IPI monitoring can identify patients with persistent thrombotic risk and whether incorporating the IPI into validated ACS risk models enhances individualized management.

**Highlights:**

- We derived an Integrated Platelet Index (IPI) representing a PCA-based morphological phenotype from five routine platelet indices.
- Lower IPI values identified patients with heightened long-term atherothrombotic risk after acute coronary syndrome.
- The IPI captures multidimensional platelet alterations shaped by thrombo-inflammatory and cardiometabolic stress.
- The association between IPI and adverse outcomes remained consistent across major cardiometabolic subgroups.
- The IPI provides an accessible, low-cost phenotype with potential utility for risk stratification in secondary prevention.

## Introduction

Cardiovascular disease remains the leading cause of global mortality, and acute coronary syndrome (ACS) continues to impose a substantial clinical and economic burden[1]. Platelets play a pivotal role in atherothrombosis and contribute to inflammatory and immune pathways that underpin atherosclerotic progression and plaque destabilization[2, 3]. Increasing evidence indicates that platelet morphology and turnover are influenced by cardiometabolic disturbances—including insulin resistance, dyslipidemia, obesity, and chronic low-grade inflammation—which collectively shape a thrombo-inflammatory milieu and may predispose ACS patients to recurrent adverse events[4]. However, despite their biological relevance, routinely reported platelet indices have not been effectively incorporated into clinical risk assessment.

Commonly available indices—platelet count (PLT), mean platelet volume (MPV), platelet distribution width (PDW), plateletcrit (PCT), and platelet large-cell ratio (P-LCR)—offer complementary information regarding platelet abundance, size, heterogeneity, and circulating mass. Yet each parameter captures only one dimension of platelet morphology or turnover, and prior studies evaluating them individually have yielded heterogeneous or conflicting associations with cardiovascular outcomes[5–8]. Because these indices are intercorrelated and influenced by shared biological processes, conventional single-parameter or multivariable approaches are limited in their ability to identify coherent platelet phenotypes relevant to thrombotic risk.

Principal component analysis (PCA) provides a framework for integrating correlated hematologic markers into composite variables that reflect underlying biological patterns rather than isolated parameters[9]. Leveraging this approach, we derived an Integrated Platelet Index (IPI) from five routinely measured platelet indices—PLT, MPV, PDW, PCT, and P-LCR—to characterize a multidimensional platelet morphological phenotype potentially indicative of enhanced platelet turnover and prothrombotic propensity. Because platelet morphology is shaped by metabolic and inflammatory stressors common in ACS, such an integrated phenotype may hold particular relevance for long-term atherothrombotic risk. The objectives of this study were to (1) characterize the association between the PCA-derived Integrated Platelet Index (IPI) and long-term atherothrombotic outcomes in ACS, and (2) examine the consistency of this platelet morphological phenotype across cardiometabolic conditions known to modulate thrombo-inflammatory pathways.

## Methods

### Study Population

All patients in this prospective cohort study were derived from the Beijing Hospital Atherosclerosis Study (BHAS) (ClinicalTrials.gov identifier: NCT03072797). The study consecutively enrolled patients who underwent coronary angiography at Beijing Hospital between March 2017 and October 2020. Eligible participants were adults who were either suspected of having coronary artery disease (CAD) or had a confirmed history of CAD. Baseline demographic characteristics (including age and sex), medical history (including diabetes mellitus, hypertension, and dyslipidemia), cigarette smoking status, and body mass index (BMI) were systematically recorded. Venous blood samples were collected within 24 hours of hospital admission and prior to coronary angiography, and all laboratory parameters were measured using standardized procedures. The study protocol was approved by the Ethics Committee of Beijing Hospital (Approval No. 2016BJYYEC-121-03), and written informed consent was obtained from all participants prior to enrollment.

A total of 2,970 patients were initially enrolled. Patients with missing platelet parameters or incomplete follow-up were excluded from the final analysis, and no imputation was performed. After excluding 1,208 patients without an ACS diagnosis, the cohort comprised 1,762 confirmed ACS patients. A further 260 patients were excluded due to missing platelet parameters or incomplete follow-up, resulting in a final analytic cohort of 1,467 eligible ACS patients. Participants were categorized into event and event-free groups based on major adverse cardiovascular and cerebrovascular events (MACCE) occurrence during follow-up (see Figure S1).

### Variable Definitions

The diagnosis of ACS adhered to the 2023 European Society of Cardiology (ESC) guidelines for managing acute coronary syndromes[10]. Coronary lesion complexity was assessed using the Gensini score, as previously described[11].

### Principal Component Analysis (PCA)

The principal components (PCs) are a new set of variables organized according to their capacity to describe a given phenomenon. In order to calculate the PCs from an original data matrix X, it is necessary to eliminate data heterogeneity. This can be achieved by centring the mean at zero and the variance at one for each variable in X using the equation below:

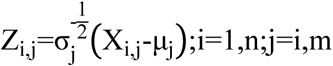

Where X_i, j_ is the i^th^ observation of variable j, Z_i, j_ is the standardized variable of X_i, j,_ µ_j_ is the mean value of variable j, and σ_j_ is the variance of variable j. Both n and m represent the total number of observations and variables, respectively.

Once the matrix Z has been obtained, the variance–covariance matrix can be derived. The PCs can be generated by calculating the eigenvector and the eigenvalues for matrix Z, and they are a linear combination of the original standardized variables:

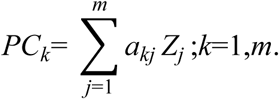

In this study, PCA was used to integrate five routine platelet parameters: PLT, MPV, PDW, PCT and P-LCR. The IPI was calculated as a composite score using the ‘linear combination coefficient matrix’ derived from the PCs. PCA extracted two PCs with eigenvalues greater than one, which accounted for 57.995% and 21.357% of the total variance, respectively (cumulative variance explained: 79.352%). The variance explanation rates and factor loadings of the PCA can be found in Additional Table S1. The IPI was calculated as a weighted sum based on the proportion of variance explained: IPI = (57.995 × Component Score 1 + 21.357× Component Score 2) / 79.352. The component score equations were:

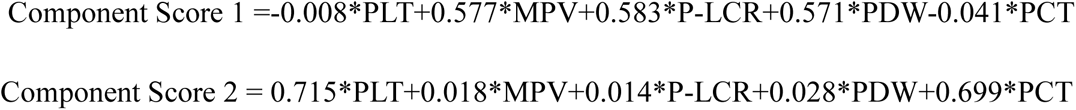

### Outcomes and Follow-up

The primary endpoint was a composite measure of MACCE, which included cardiac death, non-fatal myocardial infarction, and stroke.

### Laboratory assays

Blood routine indexes were detected using an automatic blood routine analyzer (Sysmex, Japan). Fasting blood glucose (FBG), total cholesterol (TC), triglycerides (TG), high-density lipoprotein cholesterol (HDL-C), low-density lipoprotein cholesterol (LDL-C), creatinine, Cardiac troponin I (cTnI) and Cardiac troponin T (cTnT) levels in serum samples were measured using assay kits specific to each parameter (Sekisui Medical Technologies, Osaka, Japan) on a chemical analyzer (7180 series; Hitachi, Tokyo, Japan).

### Statistical Analysis

The normal distribution of the samples was tested. Continuous variables were presented as the mean ± standard deviation (for normally distributed data) or the median (with interquartile range) (for skewed data) and were compared using independent samples *t*-tests or Mann-Whitney U tests. Categorical variables were presented as frequencies (%), compared using Pearson’s chi-square test. To minimize potential sources of bias, the prognostic value of the IPI was assessed using multivariable Cox regression implemented in four sequential models. Model 1 was unadjusted, providing the crude association. Model 2 was adjusted for basic demographic characteristics, including sex and age. Model 3 further included variables that demonstrated statistical significance (p < 0.05) in the univariate Cox regression analysis, thereby accounting for key clinical confounders. Finally, Model 4 represented the most comprehensive adjustment, building upon Model 3 by incorporating additional variables of smoking status, coronary artery type, hypersensitive C-reactive protein, cardiac troponin I, and heart rate to provide a fully adjusted hazard ratio. Kaplan-Meier analysis was used to compare survival rates between ACS patients who were stratified into IPI tertiles, and differences were assessed using log-rank tests. Threshold effect analysis explored potential inflection points in the IPI-MACCE relationship. The potential non-linear relationship between the IPI and clinical endpoints was assessed using restricted cubic splines (RCS), with knot locations optimized using the Akaike information criterion. A robustness test was performed to evaluate the consistency of the IPI’s prognostic value. This involved applying the optimal IPI cut-off value of 0.094, identified from the receiver operating characteristic (ROC) curve, alongside other grouping methodologies. Data processing and analysis were performed using R version 4.3.3 (29 February 2024), along with Zstats 1.0 (www.zstats.net).

## Results

### Baseline Characteristics

A total of 1,467 patients with ACS were included in the final analysis of this study(Figure 1). The median follow-up duration was 55.0 months (interquartile range: 40.5–70.0 months). During this period, 141 patients (9.6%) experienced the occurrence of MACCE. The baseline demographic, clinical, and laboratory characteristics of the total population and the two groups stratified by MACCE occurrence are detailed in Table 1.

**Figure 1.**
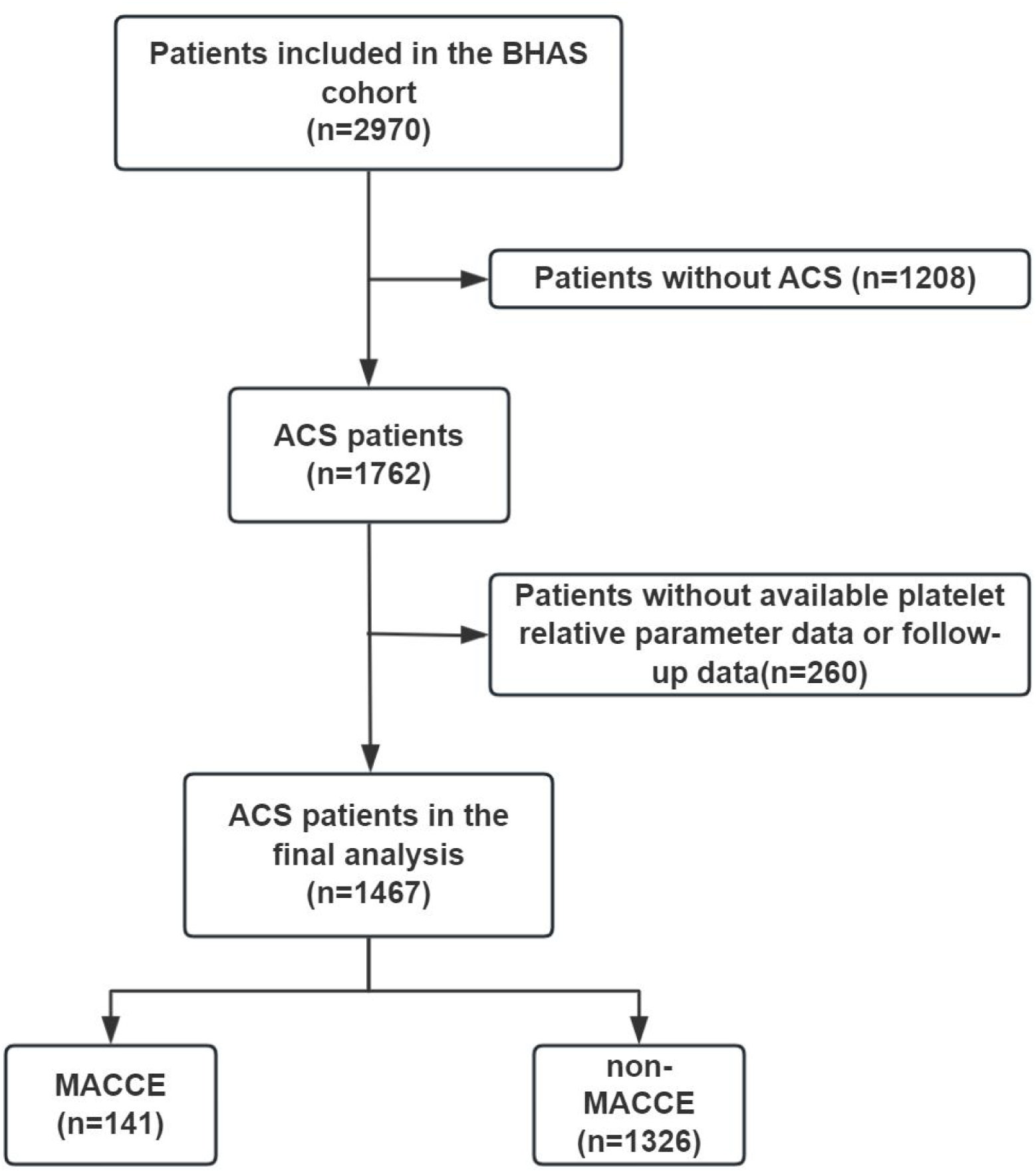
Patient enrollment flowchart. BHAS, Beijing Hospital Astherosclerosis Study; ACS, acute coronary syndrome; MACCE, major adverse cardiovascular and cerebrovascular events.

**Table 1.**
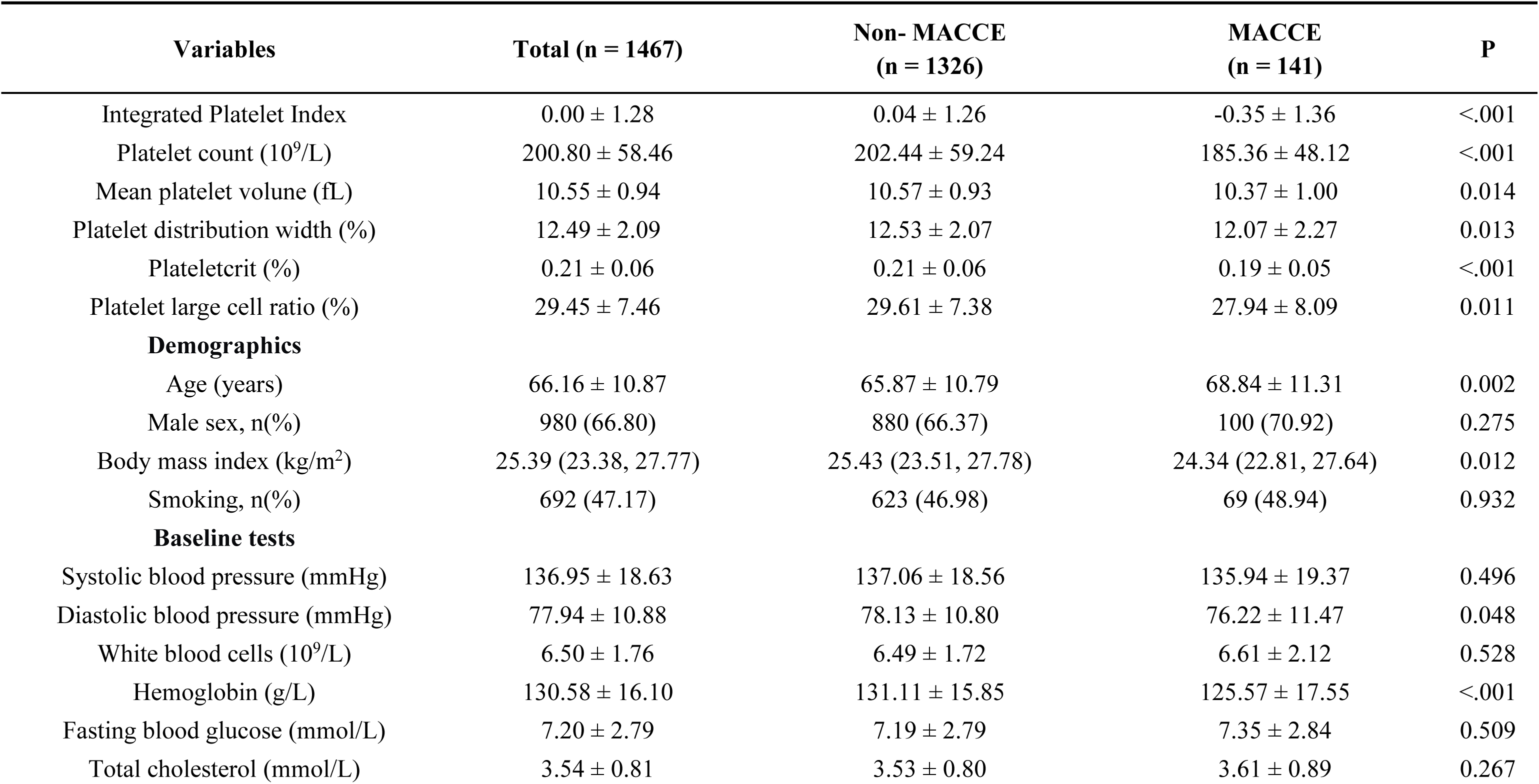
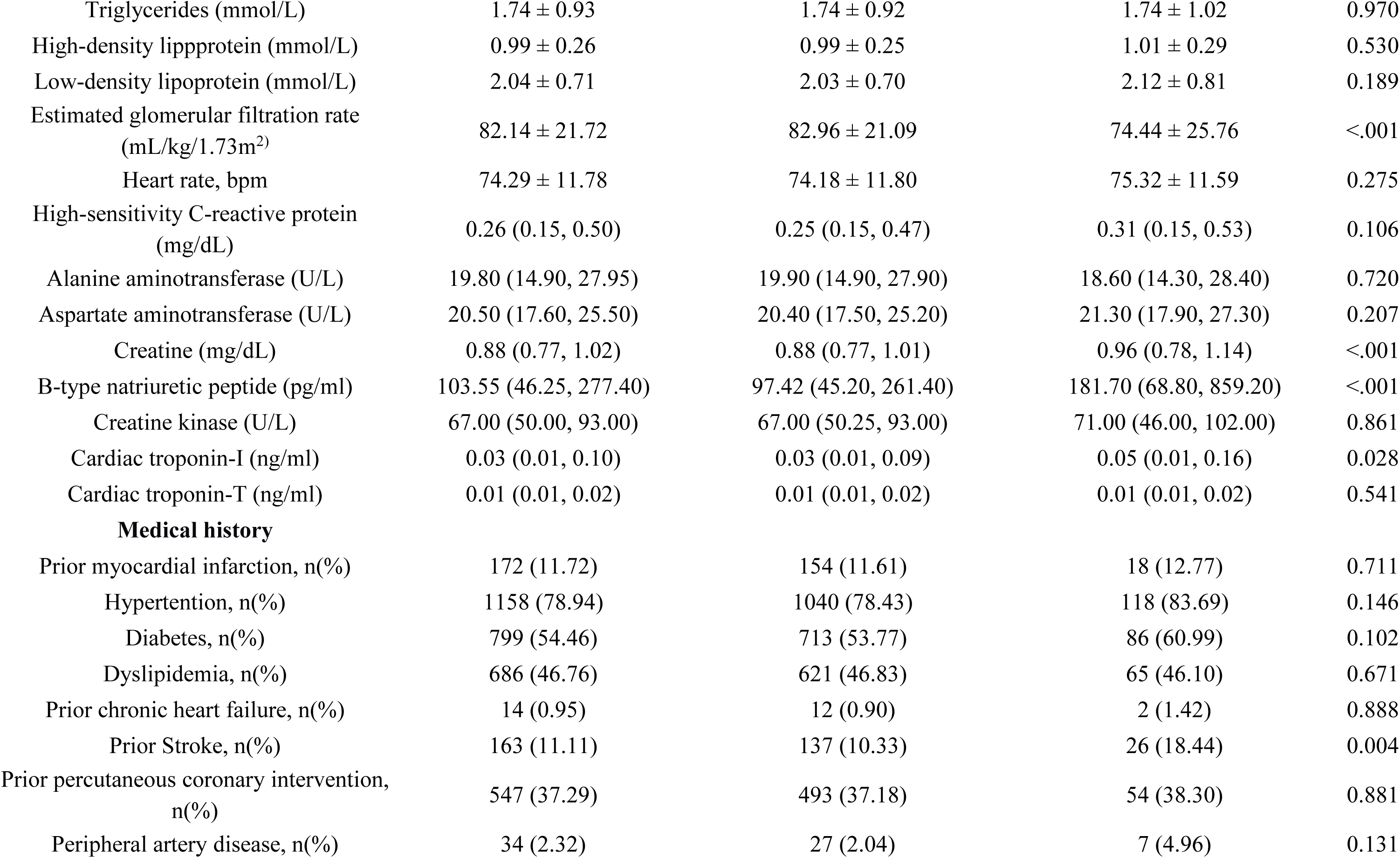
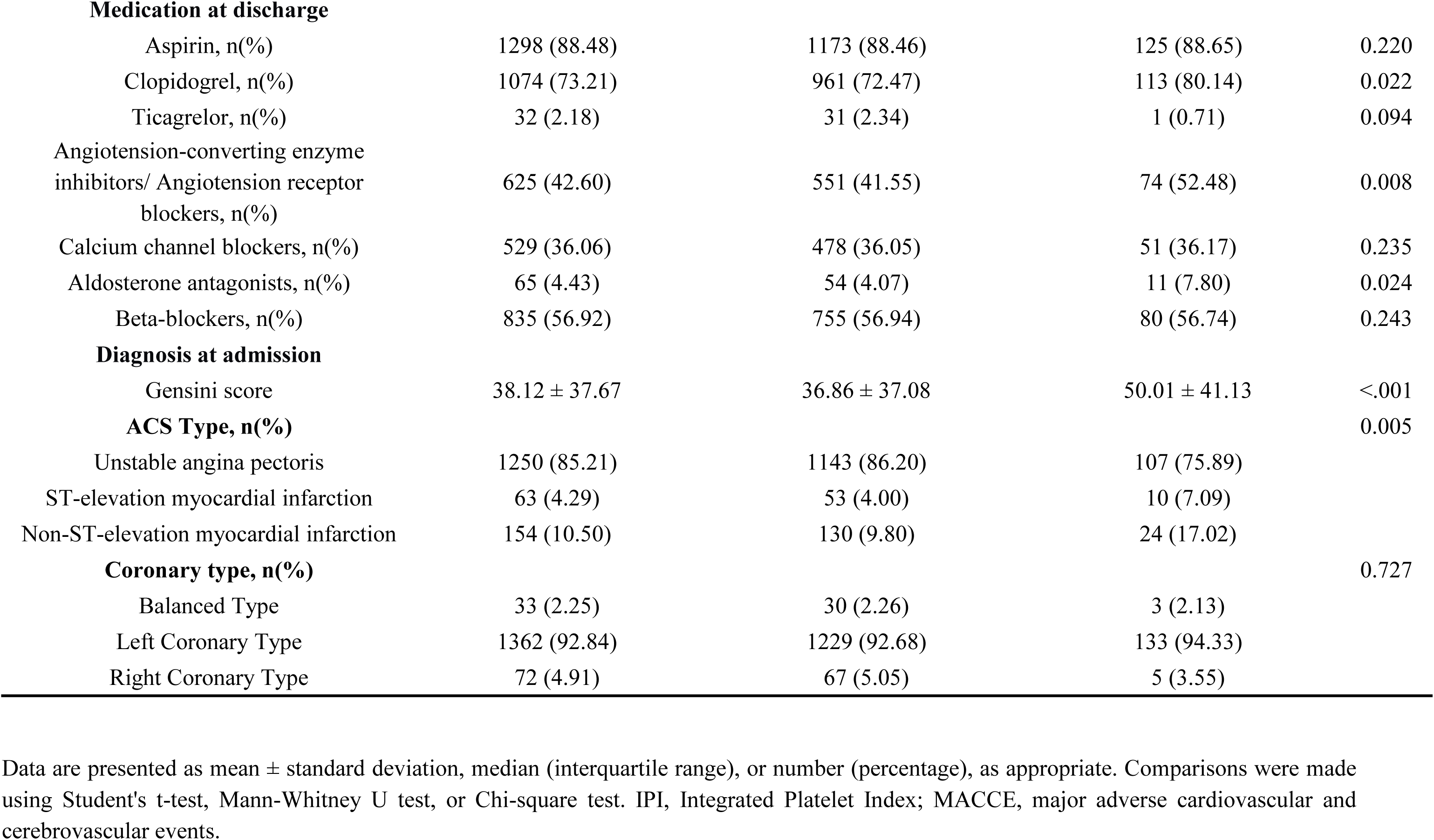
Baseline Clinical and Laboratory Characteristics of the Study Population.

At baseline, the mean age of the cohort was 66.16 ± 10.87 years, and 980 patients (66.8%) were male. Patients in the MACCE group were significantly older and had a lower body mass index (BMI), and they showed trends toward a higher prevalence of diabetes, although the difference was not statistically significant. They also exhibited lower levels of diastolic blood pressure, hemoglobin, and estimated glomerular filtration rate (eGFR) levels, as well as higher creatinine, B-type natriuretic peptide (BNP), and Gensini score levels, indicating greater coronary artery disease severity. Furthermore, there were significant differences between the two groups in all five individual platelet parameters(PLT, PCT, MPV, P-LCR, and PDW)(all P < 0.05).

### IPI Construction

Before performing PCA, all variables were standardized using z-score normalization to ensure comparability across different units. The suitability of the data for PCA was confirmed using the Kaiser-Meyer-Olkin (KMO) measure and Bartlett’s test of sphericity. The KMO value exceeded the threshold of 0.6 at 0.726, and Bartlett’s test was statistically significant (P < 0.05), both of which support the factorability of the data.

Notably, the IPI was significantly lower in the MACCE group than in the non-MACCE group(-0.35 ± 1.36 vs. 0.04 ± 1.26, P < 0.001), which is consistent with the trends observed in the baseline platelet parameters.

### Association Between IPI and MACCE

Univariate Cox regression analysis identified several baseline variables that were significantly associated with MACCE. The complete univariate analysis for all variables is provided in Additional Table S2. When modelled as a continuous variable, the IPI was significantly associated with a lower risk of MACCE in univariable Cox regression analysis (HR= 0.77, 95%CI: 0.67–0.89; p < 0.001). Table 2 then shows the results after multivariable adjustment for covariates. Following full adjustment for age, clinical characteristics, and key laboratory and coronary disease severity indicators, a 27% reduction in the risk of MACCE was independently associated with each unit increase in the IPI(aHR = 0.73, 95% CI: 0.59–0.90) (see Table 2).

**Table 2.** Unadjusted and adjusted associations between the IPI and MACCE.

| Variables | Model1 |  | Model2 |  | Model3 |  | Model4 |  |
| --- | --- | --- | --- | --- | --- | --- | --- | --- |
|  | HR (95%CI) | *P* | HR (95%CI) | *P* | HR (95%CI) | *P* | HR (95%CI) | *P* |
| IPI | 0.77(0.67~0.89) | <.001 | 0.77(0.67~0.89) | <.001 | 0.76(0.67~0.88) | <.001 | 0.73(0.59~0.90) | 0.003 |

**Model 1:** unadjusted
**Model 2:** Adjust for age, sex.
**Model 3:** Adjust for age, red blood cells, hemoglobin, aspartate aminotransferase, uric acid, creatine, estimated glomerular filtration rate, Gensini score, albumin, B-type natriuretic peptide, creatine kinase, diabetes, prior stroke, peripheral artery disease, clopidogrel, angiotension-converting enzyme inhibitors/ angiotension receptor blockers, and acute coronary syndrome type.
**Model4:** Model3 plus smoking, coronary type, high-sensitivity C-reactive protein, cardiac troponin-I, and heart rate.
IPI, Integrated Platelet Index; MACCE, major adverse cardiovascular and cerebrovascular events; HR, Hazard Ratio; CI, Confidence Interval.

The dose-response relationship between the continuous IPI and MACCE risk was examined using RCS. This analysis identified a significant non-linear pattern (P for non-linearity = 0.036), which is visualized in Figure 2. The relationship exhibited a threshold effect: MACCE risk decreased in a dose-dependent manner as the IPI increased from low values, but this trend reached a plateau at higher IPI levels, indicating a saturation point. A specific inflection point was quantified at an IPI of 1.37. Below this value, each unit increase in IPI was associated with a reduction in MACCE risk (HR = 0.63). However, beyond this threshold, no significant association was observed (P for likelihood ratio test = 0.059; see Additional Table S3).

**Figure 2.**
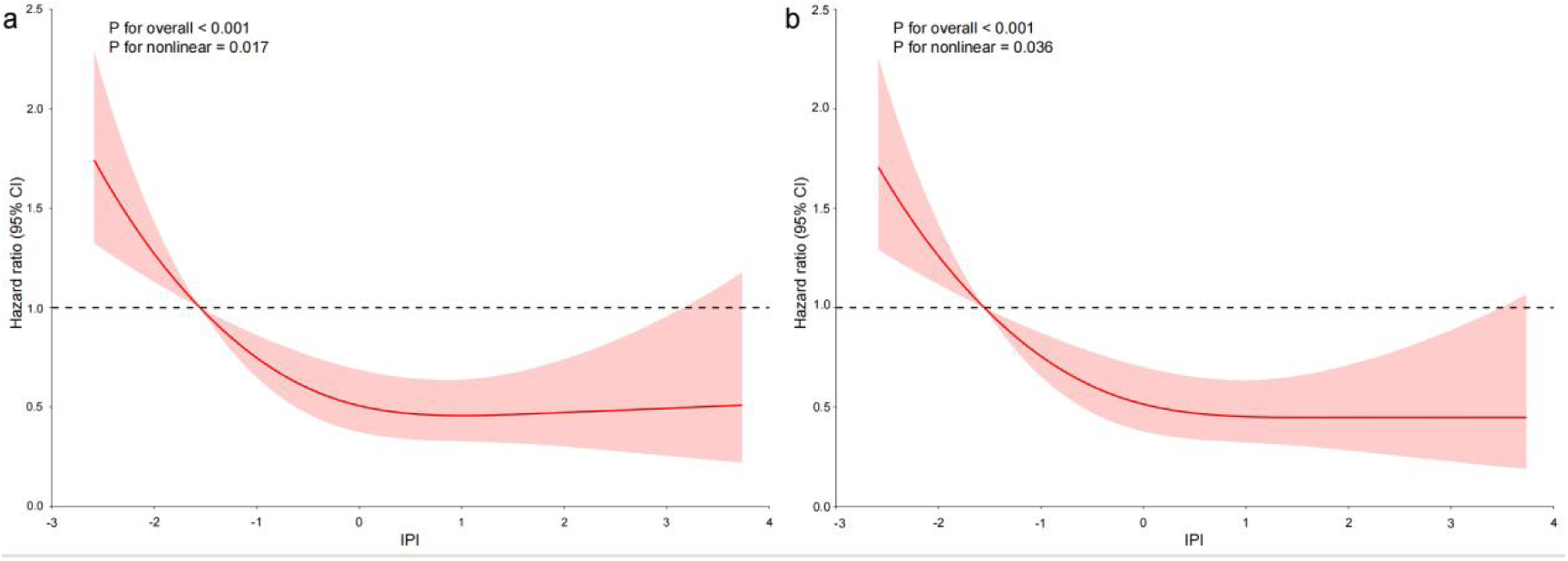
Nonlinear association between the continuous IPI and MACCE risk. a.The restricted cubic spline plot illustrates the unadjusted dose-response relationship of IPI as a continuous variable with MACCE risk, derived from a univariable Cox proportional hazards model. b. The plot illustrates the dose-response relationship after adjustment for covariates in Model 3, derived from a multivariable Cox proportional hazards model. In both panels, the solid line represents the hazard ratio (HR), and the shaded area represents the 95% confidence interval. The reference value (HR = 1) was set at the first inflection point (k1) of the RCS curve, which corresponded to the 10th percentile of the IPI distribution (IPI = -1.55). The P-value for nonlinearity was 0.036 for the overall association.

Given this non-linear relationship, we categorized patients into tertiles to visualize the difference in long-term clinical outcomes. When patients were stratified into low, middle, and high groups based on IPI tertiles, Kaplan-Meier curves demonstrated a clear gradient in event-free survival across IPI tertiles, with the highest tertile exhibiting the most favourable prognosis (log-rank P=0.006, see Figure 3). Event counts and incidence rates across IPI tertile groups are presented in Additional Table S4. A decreasing trend in the incidence of MACCE was observed from the lowest to the highest IPI tertile, accompanied by a reduction in both the number of events and person-month–adjusted incidence rates. The highest tertile showed the lowest incidence rate (458.33 per 1000 person-months), compared with 652.78 and 847.22 in the middle and lowest tertiles, respectively. To facilitate clinical use, we also stratified patients based on a pragmatic cut-off value. Similar survival curves were observed when patients were dichotomized using the optimal cut-off value derived from ROC curve analysis (IPI = 0.094; log-rank P < 0.001). The incidence of MACCE was significantly higher in the low-IPI cut-off group (11.86%) than in the high-IPI cut-off group (6.78%) (P < 0.001, see Additional Figure S1). A comparison of the baseline characteristics between the low- and high-IPI cut-off groups is provided in Additional Table S5. Importantly, Kaplan-Meier survival analysis was conducted using three distinct IPI categorization methods, all of which yielded highly consistent results (see Additional Figure S1).

**Figure 3.**
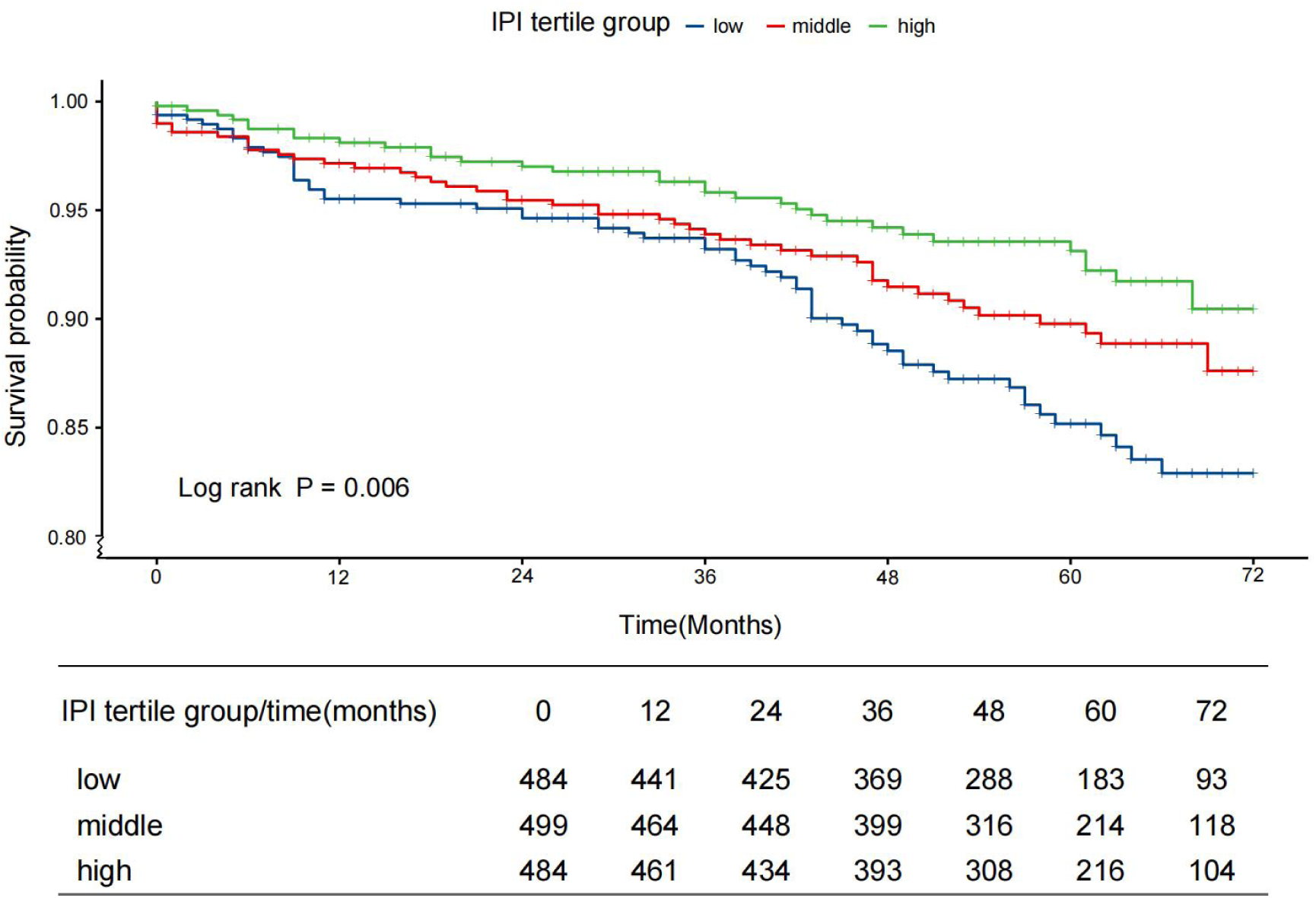
Kaplan-Meier analysis of event-free survival stratified by the IPI. Patients were categorized into low, middle, and high tertiles based on their baseline IPI value. The cumulative incidence of major adverse cardiovascular and cerebrovascular events (MACCE) was significantly lower in the highest IPI tertile compared to the lowest tertile (log-rank test, P = 0.006). IPI, integrated platelet index.

To assess the robustness of the association between the IPI and MACCE, subgroup analyses were performed across clinically relevant strata (Figure 4). The inverse association between a higher IPI and MACCE risk was consistently observed in all examined subgroups. Notably, the protective effect remained similar in both patients with and without diabetes(P for interaction = 0.935), dyslipidemia (P for interaction = 0.834), and across BMI categories (P for interaction = 0.547), indicating that neither metabolic status nor adiposity materially modified the relationship.

**Figure 4.**
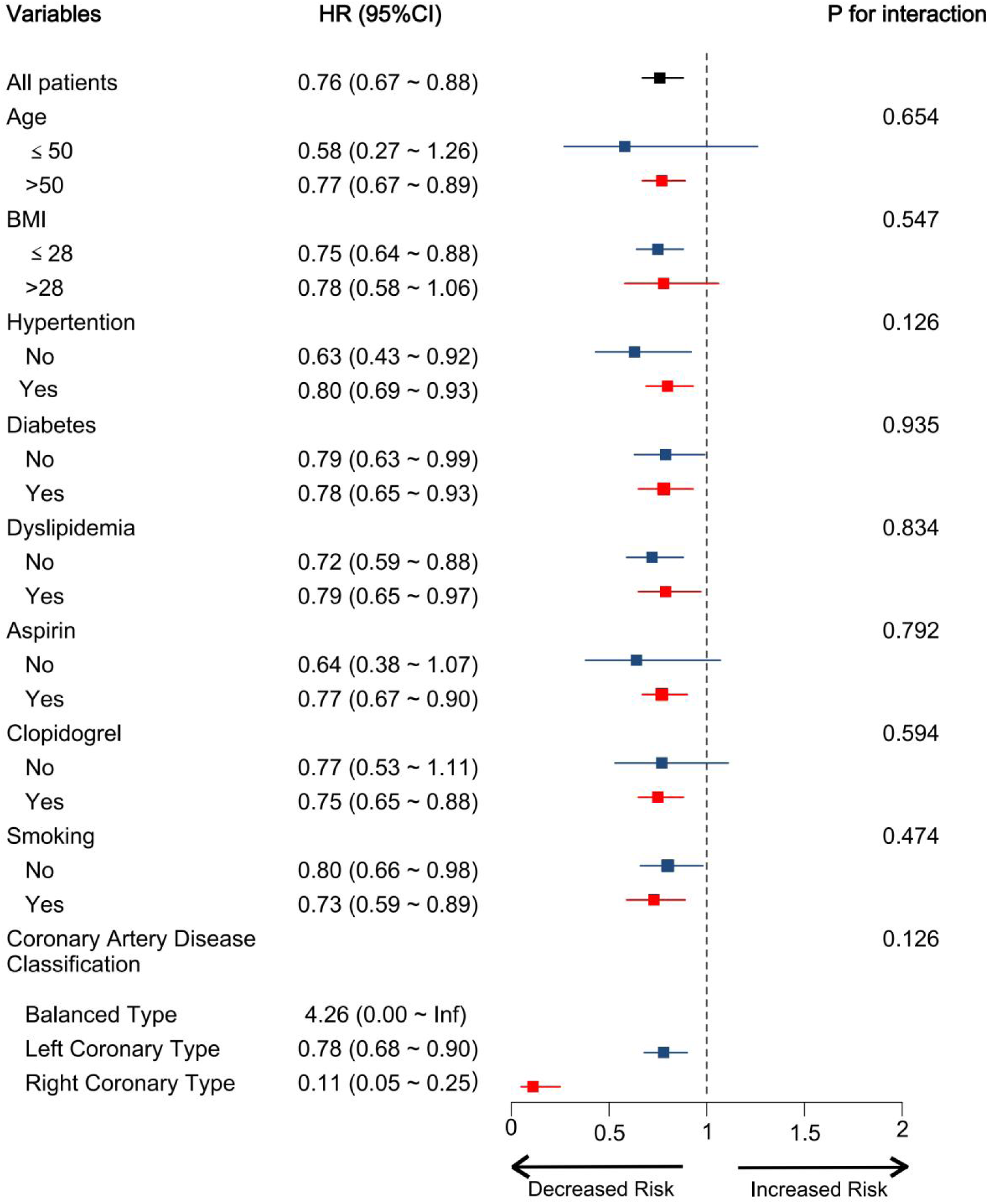
Forest plot of subgroup analyses for the association between the IPI and MACCE. Forest plot shows hazard ratios (squares) and 95% confidence intervals (horizontal lines) for the association of IPI (per 1-unit increase) with MACCE risk across patient subgroups. The size of the square is proportional to the precision of the estimate. The vertical dashed line indicates the overall hazard ratio for the entire cohort. Subgroup analyses were based on Model 3. P-values for interaction were derived from the likelihood ratio test. IPI, Integrated Platelet Index; MACCE, major adverse cardiovascular and cerebrovascular events.

## Discussion

In this prospective cohort of patients with ACS, we developed a principal-component–derived Integrated Platelet Index (IPI) that synthesizes five routinely measured platelet indices into a single morphological phenotype. We found that lower IPI values were associated with a markedly higher long-term risk of MACCE. The association was non-linear, with the steepest rise in risk observed at lower IPI values, suggesting that relatively small perturbations in platelet morphology—potentially indicative of heightened turnover or activation propensity—may carry disproportionate prognostic relevance. Although threshold analysis did not identify a statistically discrete inflection point, the smooth L-shaped curve and risk plateau at higher values support the concept of a continuous, biologically graded phenotype rather than one governed by a strict threshold. These characteristics indicate that the IPI may be particularly sensitive in identifying patients with a more adverse platelet profile and may be most clinically informative within the lower range of its distribution. Importantly, the consistent inverse association across prespecified clinical and metabolic subgroups underscores the robustness of this phenotype despite heterogeneous cardiometabolic backgrounds.

Platelets contribute not only to acute thrombus formation following plaque rupture but also to chronic inflammatory and immune processes that shape atherosclerotic disease progression[12]. Cardiometabolic disturbances—including insulin resistance, dyslipidemia, obesity, and persistent low-grade inflammation—are known to remodel platelet morphology and turnover, fostering a prothrombotic, pro-inflammatory state that may amplify risk in ACS[4, 13]. Routine complete blood counts provide readily accessible parameters that capture distinct facets of platelet biology. PLT reflects the abundance of platelets in circulation; MPV and PDW serve as proxies for platelet size and size heterogeneity, which are often linked to the activation state; and PCT and P-LCR indicate the total mass of platelets and the proportion of larger, potentially more reactive platelets, respectively. Although complete blood count–derived platelet indices capture important aspects of platelet size, heterogeneity, and circulating mass, individual parameters have shown inconsistent associations with cardiovascular outcomes. For example, while a large-scale meta-analysis confirmed that each 1 fL increase in MPV is associated with a 29% higher long-term mortality risk (HR 1.29) [14], the effect estimates for MPV vary widely across studies (HR: 0.90 to 1.66)[15], indicating inconsistent predictive strength. Similarly, the relationship between PLT and outcomes is U-shaped rather than linear, with both thrombocytopenia and thrombocytosis being linked to increased mortality[16]. Evidence for other parameters, such as PDW, remains inconclusive[6, 7, 17, 18]. However, inconsistent effect sizes, unclear cut-offs, and the unidimensional nature of each parameter restrict their clinical utility. These inconsistencies likely reflect the unidimensional nature of individual parameters, their intercorrelation, and the multifaceted biological processes influencing platelet morphology. Such limitations highlight the need for integrative phenotyping approaches capable of capturing coordinated alterations that reflect the broader thrombo-inflammatory milieu.

PCA addresses these challenges by integrating correlated variables into components that represent shared biological variance rather than isolated traits[19–24]. In our study, PCA condensed five platelet indices into a morphological phenotype—quantified as the IPI—that may reflect platelet turnover, size distribution, and heterogeneity, all of which are linked to metabolic and inflammatory stress. The consistency of the IPI–MACCE association across modeling approaches strengthens the validity of this composite phenotype. Notably, the finding that higher IPI values were associated with lower risk contrasts with prior studies in which elevated MPV or PDW individually appeared to predict worse outcomes. This divergence suggests that composite phenotyping may offer a more accurate representation of the platelet milieu than any single parameter, which may otherwise exert competing or compensatory effects. In biological terms, a higher IPI likely reflects a platelet population characterized by more stable morphology and reduced prothrombotic propensity, whereas a lower IPI may mark increased platelet heterogeneity, higher turnover, or subtle metabolic dysregulation predisposing to adverse events.

Our findings also align with mechanistic evidence demonstrating that cardiometabolic stressors remodel platelet morphology toward larger, more heterogeneous, and more reactive forms[4, 13, 25–29]. This provides a plausible pathway through which metabolic conditions may influence the platelet phenotype captured by the IPI. At the same time, the lack of significant interaction across metabolic subgroups suggests that while metabolic stress may contribute to shaping platelet morphology, it does not materially alter the prognostic relevance of the IPI. This stability across diverse cardiometabolic backgrounds enhances its translational potential.

Our study has several strengths. First, the association between the IPI and reduced risk remained statistically significant after sequential adjustment for demographics, traditional risk factors, and potent prognostic indicators such as cardiac biomarkers, angiographic severity, and medications. Importantly, this association also persisted after accounting for metabolic comorbidities, suggesting that the IPI provides prognostic value beyond established cardiometabolic risk factors. IPI captures platelet alterations driven by metabolic stress, which traditional predictors do not quantify. Methodologically, the application of PCA enabled us to efficiently combine multiple correlated platelet parameters into a single, practical clinical index. This approach addresses the challenge of collinearity and simplifies the interpretation of multifaceted platelet physiology. It demonstrates a feasible strategy for extracting novel insights from routine laboratory data.

The IPI shows promise in enhancing the risk stratification of patients with ACS. As it is derived from standard, automated complete blood count parameters, the IPI offers several practical advantages, including low cost, widespread availability, and quick results. These characteristics make it feasible for use in a range of clinical settings, including those with limited resources. Additionally, repeated IPI measurement during follow-up could potentially serve as a dynamic biomarker reflecting an individual’s response to antiplatelet therapy and indicating residual ischaemic risk. Because platelet reactivity and turnover are strongly influenced by metabolic status, serial IPI assessment may help identify patients whose cardiometabolic milieu continues to promote thrombotic risk despite guideline-directed therapy. This could help to guide treatment intensification or de-escalation over time. Finally, incorporating the IPI into validated risk scores such as the Global Registry of Acute Coronary Events (GRACE) or Thrombolysis In Myocardial Infarction (TIMI) could improve prognostic accuracy further and support more personalized management of ACS.

Several limitations warrant acknowledgment. Firstly, the retrospective, observational design of this study inherently precludes definitive causal inferences. Despite extensive adjustment, residual confounding by unmeasured factors (e.g., detailed lifestyle factors and variations in undocumented medication adherence) cannot be fully excluded. Secondly, the results originate from a single-centre cohort. Although rigorous internal validation was performed, external validation in large, multicentre, independent, and prospectively designed cohorts encompassing diverse ACS subpopulations and global patient demographics is essential to confirm the generalisability and robustness of the IPI. Thirdly, the biological interpretation of the IPI is partly inferential, based on its constituent parameters. Future mechanistic studies directly correlating the IPI with functional platelet assays are required. Fourthly, the IPI was calculated at a single baseline time point. Assessing its trajectory during the acute phase of ACS and post-discharge could provide additional prognostic insights. Finally, the focus on the composite score meant that the relative contribution of individual platelet parameters to the IPI (PCA loadings) was not discussed in detail in this context. Such an analysis might offer a deeper biological understanding.

## Conclusions

The IPI was associated with long-term MACCE in patients with ACS. Its nonlinear risk pattern and stability across metabolic subgroups suggest that the IPI reflects integrated platelet alterations linked to cardiometabolic and inflammatory stress. As a simple composite measure derived from routine testing, the IPI may enhance risk stratification in ACS. Further validation and mechanistic investigation are needed to confirm its clinical relevance.

## Data Availability

Part data generated or analysed during this study are included in this published article [and its supplementary information files] The clinical data used in this study contain sensitive personal health information and cannot be shared publicly due to privacy and ethical restrictions. De-identified analytical datasets and statistical code used in the analysis are available from the corresponding authors upon reasonable request. Requests may be directed to Prof. Xue Yu or Prof. Wei Gong.

## Acknowledgements

The authors thank the staff of Beijing Hospital Atherosclerosis Study (BHAS) for their support in participant recruitment and data management.

No professional writing assistance was used in the preparation of this manuscript.

## Sources of Funding

This work was supported by Capital’s Funds for Health Improvement and Research (2024-1-405), National Science and Technology Major Project for the Prevention and Treatment of Cancer, Cardiovascular and Cerebrovascular, Respiratory, and Metabolic Diseases (2025ZD0546402), National High Level Hospital Clinical Research Funding (Grant BJ-2025-117; No BJ-2024-140; BJ-2025-124; BJ-2025-121),and National Natural Science Foundation of China (grant numbers 82370338).

## Disclosures

The authors declare that they have no competing interests.

## Authors’ Contributions

YW, JS, ZZ, YL, and WG contributed to the conception and design of the study.

YW, RY, JD, XJ, XC, and YLiu contributed to data acquisition, analysis, or interpretation.

YW drafted the initial manuscript.

RY, WG, and XY critically revised the manuscript for important intellectual content.

XY and WG supervised the study and contributed to funding acquisition and overall project oversight.

All authors read and approved the final manuscript and agree to be accountable for all aspects of the work.

## Notes

### Competing Interest Statement

The authors have declared no competing interest.

### Clinical Trial

NCT03072797

### Author Declarations

the Ethics Committee of Beijing Hospital (Approval No. 2016BJYYEC-121-03)

